# Thermoregulatory Stress and the Ageing Mind: Investigating Environmental High Heat Exposure as a Risk Factor for Dementia

**DOI:** 10.1101/2025.10.23.25337680

**Authors:** AT Mosaad, LA Geer, PB Barr, JL Meyers, EP Purchase-Helzner

## Abstract

**Background:** Extreme heat exposure is a growing public health threat, particularly for older adults. While air pollution is a recognized modifiable risk factor for dementia, the role of sustained high heat exposure remains underexplored. This study investigates the association between cumulative extreme heat exposure and the prevalence of Alzheimer’s disease and related dementias (ADRD) and mild cognitive impairment (MCI) in adults.

**Methods:** Data were drawn from the All of Us Research Program (n = 286 767). High heat exposure was calculated using the CDC’s National Environmental Public Health Tracking Network based on the number of extreme heat days within multi-day events from 2019 to 2023. Exposure was measured using daily maximum temperature and heat index. Diagnoses were identified from electronic health records. Logistic regression models estimated associations between heat exposure and ADRD/MCI, adjusting for demographic, health, behavioral, and socioeconomic variables. Stratified and interaction models assessed differences by age and sex.

**Findings:** Every 10 extreme heat days were associated with a 4·0% increase in ADRD/MCI odds using maximum temperature (OR=1·040, 95% CI: 1·026–1·051) and a 6·0% increase using heat index (OR=1·058, 95% CI: 1·044–1·071). Using age-stratified analysis the effect was stronger among adults aged ≥70, with odds increasing by 7·8% versus 3·9% among those <70 using heat index. A significant interaction by age was observed (p<0·001). Odds ratios were slightly higher in females, but sex did not significantly modify the association. Area deprivation accounted for 8·6%–9·6% of the association, while education and social isolation were not significant mediators.

**Interpretation:** Cumulative high heat exposure is associated with increased odds of ADRD/MCI, especially among adults aged 70 and older. These findings underscore the need for climate adaptation strategies targeting older and socioeconomically disadvantaged populations.

**Funding:** This research was supported by the All of Us Research Program, and funded by the National Institutes of Health.

**Research in context:** *Evidence before this study:* We searched PubMed and Google Scholar for studies published between January 2005 and April 2025 using combinations of the terms “extreme heat,” “heat exposure,” “heat waves,” “dementia,” “Alzheimer’s disease,” “cognitive impairment,” and “older adults.” While many studies have shown that older adults face increased mortality and morbidity during extreme heat events, few have explored cognitive outcomes. Research specifically linking heat exposure to Alzheimer’s disease and related dementias (ADRD) is scarce. Most existing studies focus on short-term heat events and general health or mortality outcomes, rather than dementia. Although some evidence suggests temporary cognitive effects from heat, robust studies on diagnosed ADRD are lacking. Only one study has assessed the impact of cumulative extreme heat on cognitive function, and it did not examine clinical diagnoses or adjust for comorbid conditions.

*Added value of this study:* To our knowledge, this is the first national-level study in the United States to examine the association between cumulative high heat exposure and diagnosed ADRD or mild cognitive impairment (MCI) using detailed individual-level electronic health records. Our study used five years of environmental exposure data (2019–2023) and two distinct metrics—maximum temperature and maximum heat index—linked to a large, diverse cohort from the All of Us Research Program. We adjusted for demographic, clinical, behavioral, and social factors and explored interactions by age and sex. We also assessed mediation of the relationship by area deprivation, education, and social isolation. Our findings reveal that cumulative heat exposure is associated with increased odds of ADRD/MCI and that risk is more pronounced among individuals aged 70 and older.

*Implications of all the available evidence:* Cumulative high heat exposure may contribute to neurocognitive risk in later life, particularly among older adults. Our findings suggest that neighborhood-level socioeconomic disadvantage partly explains this association, underscoring the influence of structural and environmental conditions on dementia risk. This research supports the integration of heat-related vulnerability into dementia prevention frameworks and climate adaptation strategies. As climate change intensifies, understanding how environmental stressors such as heat affect cognitive ageing is essential for public health planning and policy. Targeted interventions and protective measures for older adults may help mitigate these long-term cognitive impacts.

## Introduction

Extreme heat has emerged as a critical and growing public health concern. Older adults face heightened vulnerability to heat-related illness, hospitalisation, and death due to age-related physiological decline, comorbidities, and reduced thermoregulatory capacity.^1–5^ Importantly, these problems are worsening due to the accelerating pace of climate change. While the physical health consequences of extreme heat have been well-documented, its potential impact on neurocognitive health remains underexplored.

Dementia, including Alzheimer’s disease and related dementias (ADRD), is a leading cause of disability and dependency among older persons worldwide. An estimated 55 million individuals were living with dementia in 2019, a number projected to rise to 78 million by 2030 and 139 million by 2050.^6^ Dementia ranks among the top global causes of disability-adjusted life years (DALYs), with a substantial burden that has more than doubled over the past two decades; women experience approximately 60% more dementia-related DALYs than men.^7^ In the United States, Alzheimer’s disease rose from the 12th most burdensome disease in 1990 to the sixth in 2016, underscoring the rapid public health impact.^7^ These trends highlight the growing challenge dementia poses to individuals, families, caregivers, and communities. In the absence of effective disease-modifying treatments, identifying modifiable risk factors is essential to reducing the future burden of ADRD.^6,8–9^

Although several modifiable risk factors for ADRD have been identified—including high body-mass index, low education, air pollution, and cardiometabolic conditions—few studies have examined whether long-term exposure to high ambient heat contributes to dementia risk.^8–10^ Most prior studies have focused on short-term heat events or mortality outcomes, rather than the prevalence of dementia due to cumulative exposure. Few have incorporated robust adjustment for socioeconomic status, health conditions, or psychosocial stressors such as social isolation.

Given that heat exposure and dementia risk are both projected to rise steeply in the coming decades, particularly in ageing populations, there is a pressing need to understand the extent to which extreme heat contributes to ADRD. This study examines the association between cumulative high heat exposure and the prevalence of ADRD and mild cognitive impairment (MCI) in a large, diverse national sample.

## Methods

### Study design and data source

We conducted a cross-sectional study using data from Version 7 of the All of Us Research Program.^11^ All of Us is a prospective, nationwide cohort study that aims to examine the effects of environment, lifestyle, and genomic factors on health outcomes. Participant recruitment is conducted through participating health care provider organizations and in partnership with Federally Qualified Health Centers. After enrollment, participants are invited to undergo a basic physical exam and biospecimen collection at an affiliated health care site. Follow-up occurs passively via linkage with electronic health records (EHRs) and through follow-up surveys.

Participants enrolled between May 2017 and July 2022 were included (N = 287 079). Data access was granted through the program’s Controlled Tier, and analyses were conducted within the secure cloud-based Researcher Workbench.

### Study population

Participants with EHR data and complete information on ZIP code, date of birth, and relevant covariates were included. Individuals residing outside the contiguous United States or missing geographic or demographic information were excluded. The final analytic sample consisted of 286 767 participants.

### Exposure assessment

High heat exposure was defined as the total number of extreme heat days occurring within multi-day heat events (two or more consecutive days) between 2019 and 2023. To account for regional climate variability, extreme heat days were identified at the county level using 90th percentile relative thresholds based on historical distributions from 1979 to 2019. Two metrics were used: (1) daily maximum temperature and (2) daily maximum heat index. Environmental data were obtained from the U.S. Centers for Disease Control and Prevention’s National Environmental Public Health Tracking Network. County-level exposures were linked to participants’ 3-digit ZIP codes using the HUD-USPS ZIP Code Crosswalk (Q1 2023).^12^ When multiple ZIP codes fell within a single county, exposure values were averaged before merging with participant records.

### Outcome definition

The primary outcome was a binary indicator of ADRD or MCI, identified using SNOMED concepts mapped from ICD-9-CM and ICD-10-CM codes. Diagnoses were grouped using phecodes and extracted from the EHR. Participants were classified as having ADRD/MCI if at least one relevant diagnostic code was recorded during the study period.

### Covariates

Models were adjusted for demographic, clinical, behavioral, and socioeconomic variables. Age was included as a categorical variable (<60, 60–69, 70–79, 80+), and stratified analyses used a binary classification (<70 vs ≥70). Sex at birth was self-reported and categorized as male or female; participants who selected another option or declined to respond were excluded from adjusted and stratified analyses. Race and ethnicity were self-reported (non-Hispanic White, non-Hispanic Black, Hispanic, Other). Smoking status was derived from the survey question, “Have you smoked at least 100 cigarettes in your entire life?”, with responses categorized as ‘yes’ or ‘no’. Social isolation was defined as living alone based on household size. Health conditions were identified from the EHR using phecodes and included cardiovascular disease, cerebrovascular disease, depression, hearing impairment, chronic obstructive pulmonary disease (COPD), type 2 diabetes, and alcohol use disorder. Educational attainment served as the individual-level proxy for socioeconomic status (categorized as low, moderate, or high). The Area Deprivation Index (ADI) was used as the community-level measure of socioeconomic disadvantage and dichotomized into high deprivation (top 15%) versus low deprivation (bottom 85%) census block groups.^13^

### Statistical analysis

Logistic regression was used to estimate the association between cumulative high heat exposure and odds of ADRD/MCI. Separate models were fitted for each exposure metric (maximum temperature and maximum heat index). Fully adjusted models included all specified covariates. Stratified models were conducted for participants aged <70 versus ≥70 years. Interaction terms were tested to assess effect modification by age, sex, educational attainment, social isolation, and ADI.

Mediation analyses were conducted using regression-based models with 1,000 bootstrap simulations to estimate the average causal mediation effect (ACME), average direct effect (ADE), total effect, and proportion mediated. Educational attainment, social isolation, and ADI were tested as potential mediators of the association between high heat exposure and ADRD/MCI. ADI was treated as a continuous variable for mediation analyses, while educational attainment and social isolation were categorical.

To examine non-linear associations between cumulative heat exposure and ADRD/MCI, we used restricted cubic spline regression with inflection points placed at the 10th, 50th, and 90th percentiles of the exposure distribution.

Missing data were handled using listwise deletion. This approach was selected due to the low proportion of missingness across key variables (≤4·1%). All analyses were conducted in R version 4.4.0.

### Ethics approval

All participants provided informed consent to participate in the All of Us Research Program. This study used de-identified data and was conducted within the secure Researcher Workbench environment. The Institutional Review Board at SUNY Downstate Health Sciences University reviewed the project and determined that it does not constitute human subjects research as defined by federal regulations. Therefore, IRB approval was not required.

## Results

### Sample characteristics

Among the 286 767 participants included in the analysis, 4 946 (1·72%) had a documented diagnosis of ADRD/MCI. The mean age of all participants was 55·1 years (SD 17·0), with 52·8% under age 60, 21·8% aged 60–69, 17·1% aged 70–79, and 6·5% aged 80 years or older.

Prevalence of ADRD/MCI was highest among participants aged 80 years and older, followed by those in the “other” sex category, males, and individuals identifying as non-Hispanic White.

Elevated rates were also observed among participants with cardiovascular disease, cerebrovascular disease, type 2 diabetes, chronic obstructive pulmonary disease (COPD), hearing impairment, alcohol use disorder, and depression. Prevalence was also greater among participants with low educational attainment, low area deprivation, and social isolation (p < 0·05 for all comparisons; Table 1).

**Table 1:**
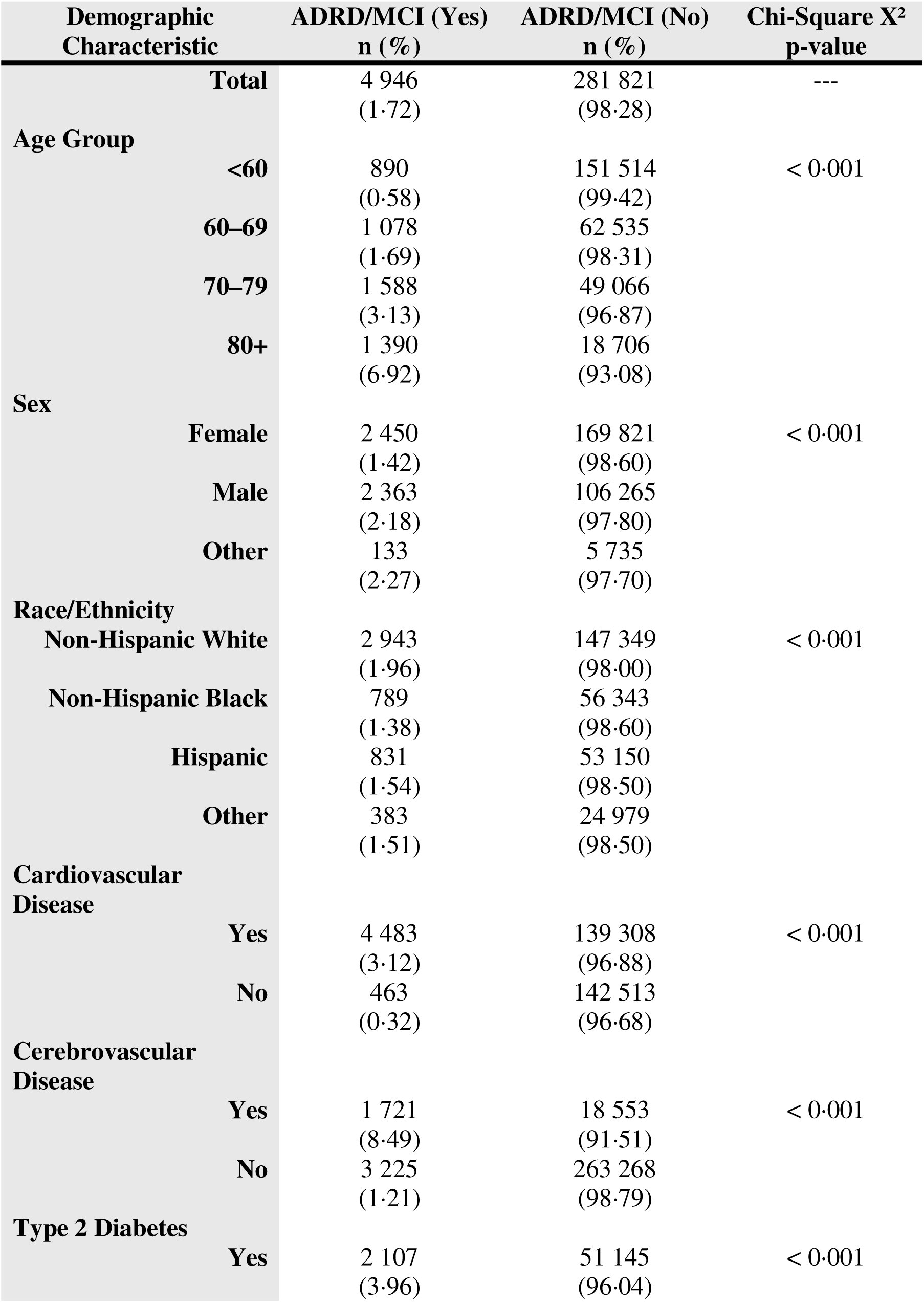
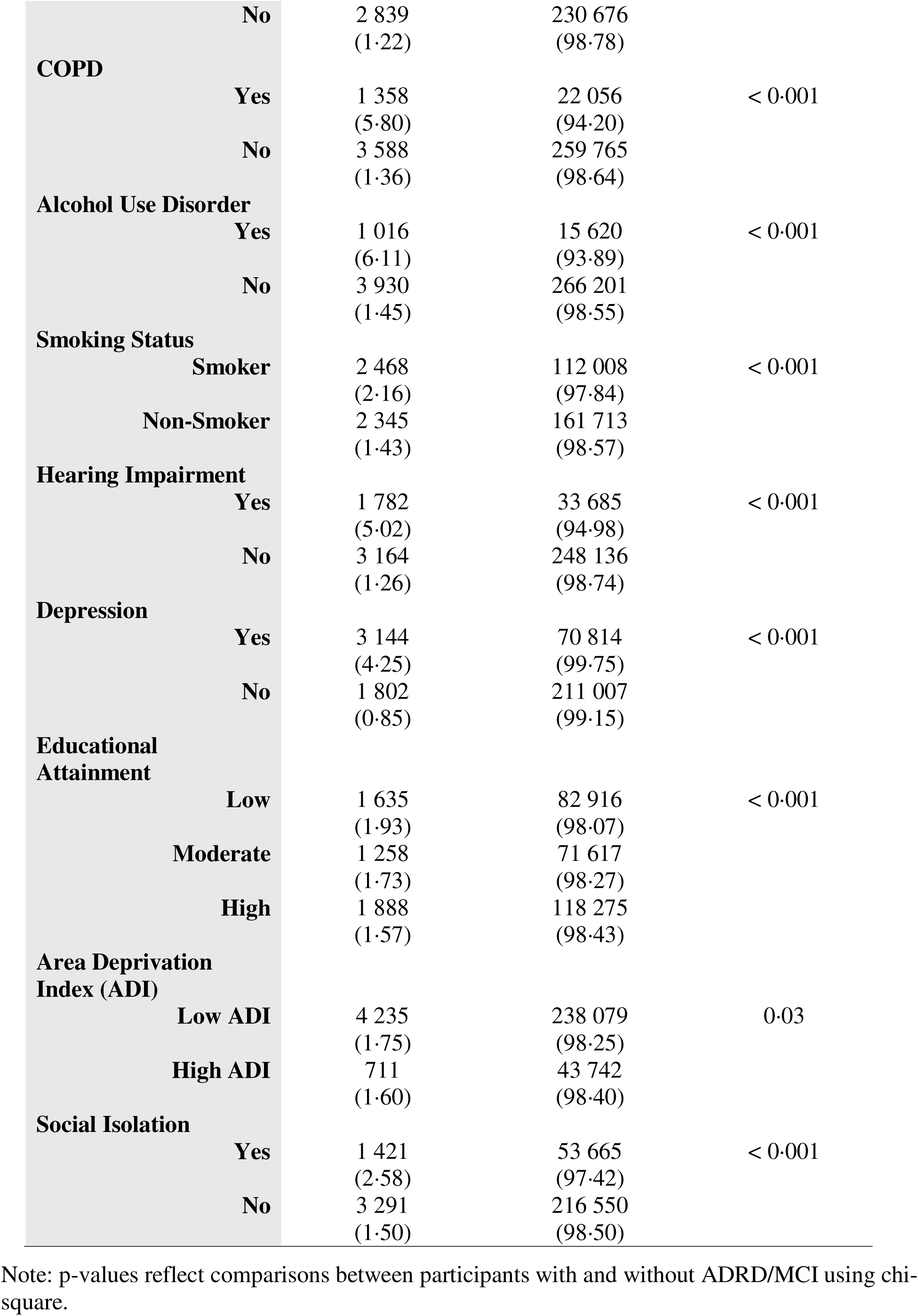
Baseline characteristics by ADRD/MCI status.

### Association between heat exposure and ADRD/MCI

In crude models, each additional extreme heat day within a multi-day event was associated with a modest but statistically significant increase in odds of ADRD/MCI. The crude odds ratio (OR) was 1·0017 (95% CI: 1·0007–1·0027) when using maximum temperature and 1·0012 (95% CI: 1·0001–1·0023) when using maximum heat index (Table 2A).

**Table 2A:** Crude Odds Ratio for the Association Between High Heat Exposure and ADRD/MCI.

| Exposure Metric | Crude OR (95% CI) |
| --- | --- |
| Per 1-day increase in high heat<br>(Maximum Temperature) | 1.0017 (95% CI: 1.0007–1.0027) |
| Per 1-day increase in high heat<br>(Heat Index) | 1.0012 (95% CI: 1.0001–1.0023) |

After adjusting for demographic, health, and socioeconomic covariates, the associations were stronger. The adjusted odds of ADRD/MCI increased by 0·39% per extreme heat day (aOR = 1·0039, 95% CI: 1·0026–1·0051) based on maximum temperature and by 0·58% (aOR = 1·0058, 95% CI: 1·0044–1·0071) based on maximum heat index (Table 2).

**Table 2:** Adjusted Odds Ratios for Heat Exposure Metrics and ADRD/MCI.

| **Covariate** | **aOR (95% CI)**<br>**Max. Temp** | **aOR (95% CI)**<br>**Heat Index** |
| --- | --- | --- |
| **Per 1-day Increase in High Heat** | 1.0039 (1.0026–1.0051) | 1.0058 (1.0044–1.0071) |
| **Age 60–69 vs <60** | 1.79 (1.62–1.97) | 1.80 (1.63–1.99) |
| **Age 70–79 vs <60** | 3.36 (3.05–3.71) | 3.40 (3.09–3.75) |
| **Age 80+ vs <60** | 7.63 (6.86–8.50) | 7.70 (6.91–8.57) |
| **Female vs Male** | 0.77 (0.72–0.82) | 0.77 (0.72–0.82) |
| **Hispanic vs White** | 1.33 (1.20–1.46) | 1.29 (1.18–1.42) |
| **Black vs non-Hispanic White** | 1.01 (0.91–1.11) | 1.00 (0.91–1.11) |
| **Other vs non-Hispanic White** | 1.20 (1.05–1.36) | 1.20 (1.05–1.36) |
| **Cardiovascular disease** | 2.77 (2.48–3.11) | 2.76 (2.47–3.09) |
| **Cerebrovascular disease** | 2.42 (2.26–2.60) | 2.43 (2.26–2.61) |
| **Type 2 diabetes** | 1.29 (1.20–1.38) | 1.28 (1.20–1.37) |
| **COPD** | 1.34 (1.24–1.45) | 1.33 (1.23–1.44) |
| **Alcohol use disorder** | 2.73 (2.50–2.98) | 2.75 (2.52–3.00) |
| **Current Smoker** | 0.90 (0.85–0.97) | 0.90 (0.85–0.97) |
| **Hearing impairment** | 1.34 (1.25–1.44) | 1.37 (1.28–1.47) |
| **Depression** | 3.43 (3.20–3.68) | 3.43 (3.20–3.68) |
| **Moderate vs High Education** | 0.98 (0.91–1.06) | 0.98 (0.90–1.06) |
| **Low vs High Education** | 1.21 (1.11–1.31) | 1.20 (1.10–1.30) |
| **High ADI vs Low ADI** | 1.17 (1.06–1.29) | 1.13 (1.02–1.24) |
| **Social isolation (Yes vs No)** | 1.05 (0.98–1.12) | 1.05 (0.98–1.12) |

### Age-stratified analyses

The association between high heat exposure and ADRD/MCI was stronger among participants aged 70 years and older. Using daily maximum temperature, each additional heat day within a multi-day heat event was associated with a 0·49% increase in the odds of ADRD/MCI among participants aged ≥70 (aOR = 1·0049, 95% CI: 1·0033–1·0065) compared to a 0·31% increase among those below the age of 70 (aOR = 1·0031, 95% CI: 1·0012–1·0050) (Table 3). Using daily maximum heat index, the association was even stronger, with a 0·75% increase in odds among participants aged ≥70 (aOR = 1·0075, 95% CI: 1·0057–1·0092), and a 0·38% increase among those aged <70 (aOR = 1·0038, 95% CI: 1·0017–1·0058) (Table 3A). A statistically significant interaction between age and high heat exposure was observed in the maximum heat index model (p < 0·001), but not in the maximum temperature model (p = 0·14).

**Table 3:** Age-Stratified Adjusted Odds Ratios for the Association Between High Heat Exposure (Maximum Temperature) and ADRD/MCI.

| **Variable** | **Age <70 aOR (95% CI)** | **Age 70+ aOR (95% CI)** |
| --- | --- | --- |
| **High heat exposure (Max Temp)** | 1.0031 (1.0012–1.0050) | 1.0049 (1.0033–1.0065) |
| **Female (vs Male)** | 0.62 (0.56–0.70) | 0.86 (0.79–0.93) |
| **Hispanic (vs Non-Hispanic White)** | 0.96 (0.83–1.10) | 1.64 (1.43–1.87) |
| **Non-Hispanic Black (vs Non-Hispanic White)** | 0.72 (0.63–0.82) | 1.29 (1.13–1.47) |
| **Other Race/Ethnicity** | 1.17 (0.96–1.40) | 1.11 (0.93–1.31) |
| **≥1 Cardiovascular Condition** | 3.22 (2.79–3.73) | 2.49 (2.11–2.96) |
| **≥1 Cerebrovascular Condition** | 2.74 (2.42–3.09) | 2.68 (2.46–2.91) |
| **Type 2 Diabetes** | 1.24 (1.12–1.38) | 1.29 (1.19–1.41) |
| **COPD** | 1.55 (1.37–1.75) | 1.35 (1.22–1.49) |
| **Hearing Impairment** | 1.57 (1.39–1.77) | 1.51 (1.39–1.65) |
| **Depression** | 3.98 (3.54–4.48) | 2.76 (2.54–3.01) |
| **Alcohol Use Disorder** | 3.33 (2.97–3.74) | 1.52 (1.31–1.75) |
| **Current Smoker** | 0.94 (0.85–1.05) | 0.91 (0.83–0.99) |
| **Moderate Education (vs High)** | 1.07 (0.94–1.21) | 0.86 (0.78–0.96) |
| **Low Education (vs High)** | 1.09 (0.96–1.24) | 1.25 (1.13–1.39) |
| **High Area Deprivation** | 1.18 (1.02–1.35) | 1.15 (1.01–1.30) |
| **Social Isolation** | 1.18 (1.05–1.31) | 1.02 (0.93–1.11) |
| **Interaction: Max. Temp × Age 70+** | --- | 1.0020 (0.999–1.0050)<sup>1</sup> |

<sup>1</sup> p = 0.14 for interaction term

**Table 3A:** Age-Stratified Adjusted Odds Ratios for the Association Between High Heat Exposure (Maximum Heat Index) and ADRD/MCI.

| Variable | Age <70 aOR (95% CI) | Age 70+ aOR (95% CI) |
| --- | --- | --- |
| **High heat exposure (Heat Index)** | 1.0038 (1.0017–1.0058) | 1.0075 (1.0057–1.0092) |
| **Female (vs Male)** | 0.63 (0.57–0.69) | 0.85 (0.78–0.93) |
| **Hispanic (vs Non-Hispanic White)** | 0.95 (0.82–1.08) | 1.58 (1.38–1.80) |
| **Non-Hispanic Black (vs Non-Hispanic White)** | 0.71 (0.62–0.81) | 1.29 (1.12–1.47) |
| **Other Race/Ethnicity** | 1.17 (0.97–1.40) | 1.11 (0.93–1.31) |
| **≥1 Cardiovascular Condition** | 3.21 (2.78–3.73) | 2.47 (2.09–2.94) |
| **≥1 Cerebrovascular Condition** | 2.74 (2.42–3.09) | 2.69 (2.47–2.92) |
| **Type 2 Diabetes** | 1.24 (1.11–1.38) | 1.29 (1.18–1.40) |
| **COPD** | 1.55 (1.37–1.75) | 1.33 (1.21–1.47) |
| **Hearing Impairment** | 1.57 (1.39–1.77) | 1.51 (1.39–1.65) |
| **Depression** | 3.98 (3.54–4.48) | 2.76 (2.54–3.01) |
| **Alcohol Use Disorder** | 3.34 (2.97–3.74) | 1.53 (1.32–1.77) |
| **Current Smoker** | 0.94 (0.85–1.05) | 0.91 (0.84–0.99) |
| **Moderate Education (vs High)** | 1.06 (0.93–1.02) | 0.86 (0.78–0.95) |
| **Low Education (vs High)** | 1.07 (0.94–1.22) | 1.24 (1.12–1.38) |
| **High Area Deprivation** | 1.16 (1.00–1.33) | 1.09 (0.96–1.24) |
| **Social Isolation** | 1.17 (1.05–1.31) | 1.03 (0.94–1.12) |
| **Interaction: Heat Index × Age 70+** | --- | 1.0048 (1.0023–1.0070)<sup>1</sup> |

<sup>1</sup> p < 0.001 for interaction term

### Interaction by sex

Sex-stratified models showed slightly higher effect estimates for females, but overlapping confidence intervals indicated no statistically significant difference. Formal interaction testing confirmed no significant sex modification in either model (temperature: p = 0·78; heat index: p = 0·55).

### Mediation analyses

The area deprivation index (ADI), reflecting community-level socioeconomic disadvantage, explained 8·6% to 9·6% of the association between high heat exposure and ADRD/MCI. In both temperature- and heat index-based models, the average direct effect remained statistically significant, suggesting an independent contribution of heat exposure beyond area-level socioeconomic status. Educational attainment (individual-level SES) and social isolation did not significantly mediate the relationship.

### Exploratory analysis: non-linear associations using spline models

Restricted cubic spline regression was used to assess the potential non-linear association between cumulative high heat exposure and odds of ADRD/MCI. Models were fitted separately using daily maximum temperature and daily maximum heat index as exposure metrics, with inflection points placed at the 10th, 50th, and 90th percentiles.

A non-linear relationship was observed for both metrics. Using maximum temperature, the odds of ADRD/MCI began to increase more rapidly beyond approximately 97 extreme heat days (Figure 1). A similar inflection point was identified around 105 extreme heat days in the heat index model (Figure 2). The slopes plateaued slightly at higher exposure levels, suggesting diminishing marginal risk beyond these thresholds. Confidence intervals widened at the extremes, reflecting reduced precision due to fewer observations with very high exposure levels.

**Figure 1.**
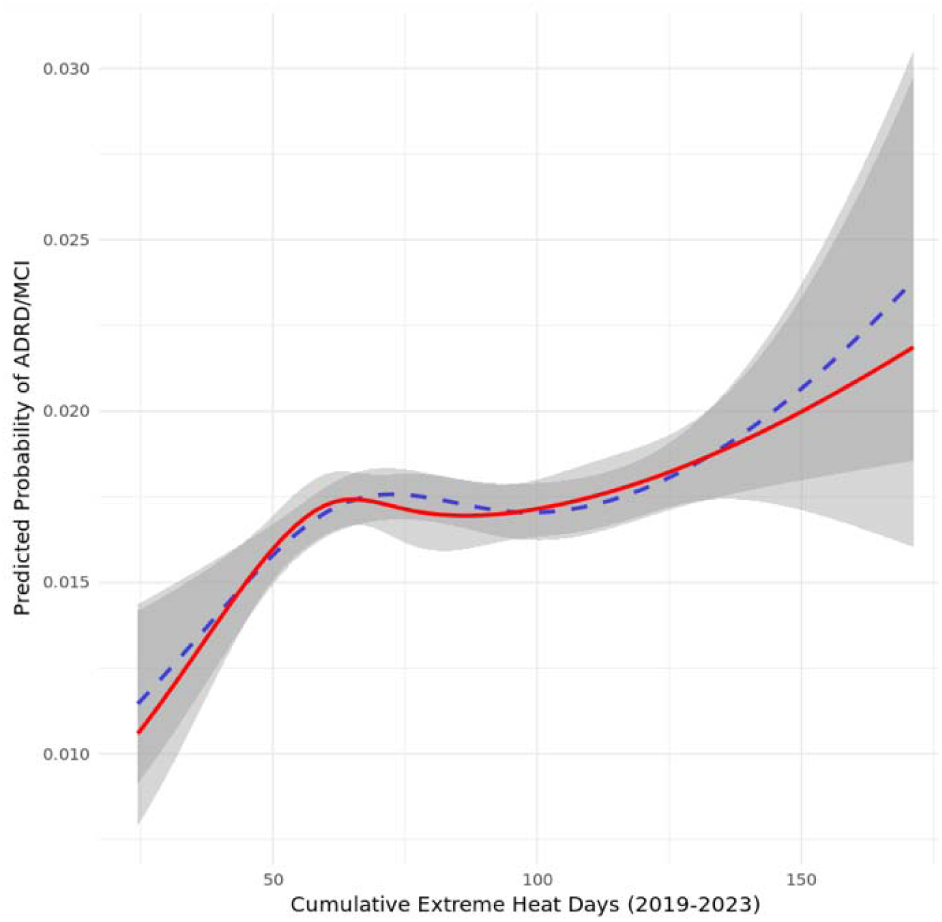
Predicted Probability Curve for ADRD/MCI Across Maximum Temperature Levels.

**Figure 2.**
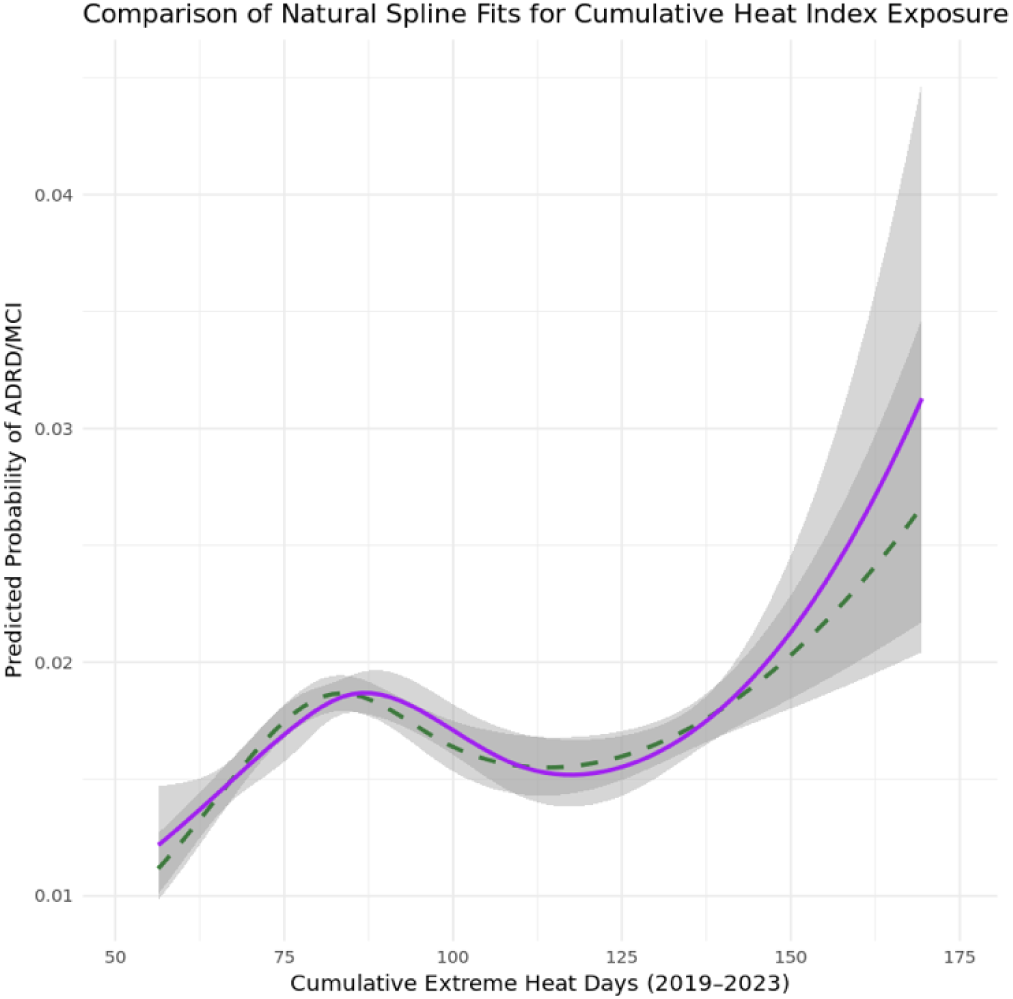
Predicted Probability Curve for ADRD/MCI Across Heat Index Levels.

## Discussion

This study adds to the growing body of evidence linking environmental stressors to cognitive decline by identifying cumulative exposure to extreme heat as a potential modifiable risk factor for Alzheimer’s disease and related dementias (ADRD) and mild cognitive impairment (MCI). Using a large, diverse, national US cohort with linked electronic health records and ZIP-code– level climate data, we found that sustained exposure to extreme heat, defined as the number of days within multi-day heat events, was associated with higher odds of ADRD/MCI. Associations were more pronounced when using maximum heat index, a composite metric that accounts for both ambient temperature and humidity, and were strongest among adults aged 70 years and older. Importantly, neighborhood-level socioeconomic disadvantage partially mediated this relationship, suggesting that social and structural factors shape vulnerability to heat-related cognitive outcomes.

These findings contribute to the evolving framework of environmental determinants of dementia. The 2020 Lancet Commission on dementia prevention, intervention, and care identified air pollution as a modifiable risk factor for dementia, alongside hypertension, low education, hearing loss, smoking, obesity, depression, physical inactivity, diabetes, and social isolation.^8^ In its 2024 update, the Commission incorporated untreated vision loss and elevated LDL cholesterol, emphasizing an increasingly holistic approach to dementia risk assessment.^10^ A recent systematic review showed that air pollution and high temperatures can interact synergistically to exacerbate health risks, including through enhanced inflammatory and cardiovascular stress responses.^14^ However, despite increasing evidence linking climate-related exposures to health outcomes, extreme heat has not yet been widely recognized as a modifiable environmental risk in dementia prevention strategies.

Our findings suggest that high heat exposure, particularly when cumulative, may warrant such consideration. The biological plausibility of this association is supported by emerging literature. Sustained exposure to elevated ambient temperatures can overwhelm thermoregulatory systems, particularly in older adults, leading to compromised cerebral perfusion, systemic inflammation, and impaired endothelial function.^15^ Experimental studies in animal models have demonstrated that chronic heat exposure increases blood–brain barrier permeability and induces oxidative stress and neuroinflammation.^15^ These processes are thought to contribute to hippocampal atrophy and neuronal dysfunction, pathological changes also observed in Alzheimer’s disease. In laboratory studies, amyloid-beta aggregation has been shown to induce intracellular thermogenesis, potentially exacerbating cellular stress and neuronal dysfunction.^16^ This is consistent with findings from a longitudinal analysis using data from the U.S. Health and Retirement Study, which found that individuals in the highest quartile of cumulative heat exposure experienced a 0·24-point greater decline in cognitive performance over six years than those in the lowest quartile.^17^ Our study builds on this evidence, demonstrating that each additional 10 days of extreme heat exposure was associated with a 4·0% to 6·0% increase in the odds of clinically diagnosed ADRD/MCI. Notably, the association reported in the study using HRS data was more pronounced among socioeconomically disadvantaged individuals and those living in deprived neighborhoods, consistent with our finding that neighborhood-level deprivation partially mediated the relationship between heat exposure and dementia risk.

Together, these findings highlight a consistent pattern across subjective cognitive decline and clinical neurocognitive outcomes, reinforcing the potential role of heat exposure in shaping late-life cognitive health.

It is important to distinguish between cumulative high environmental heat exposure and intermittent, controlled exposures such as sauna use. A prospective Finnish study in younger adults linked sauna use to reduced dementia risk, citing mechanisms such as improved vascular function, autonomic balance, and anti-inflammatory responses.^18^ These benefits likely reflect controlled, time-limited heat exposure that triggers adaptive physiological processes (hormesis), which contrasts with the sustained, uncontrolled heat exposure examined in our study. Among older adults and individuals with comorbidities, cumulative high heat may instead exacerbate vulnerability by impairing thermoregulation, increasing cardiovascular strain, and promoting systemic and neurological stress responses.^19^

We found that the association between high heat exposure and ADRD/MCI was more pronounced among participants aged 70 years and older. This finding aligns with prior work showing that older adults are biologically less able to maintain thermal equilibrium due to reduced sweat gland function, decreased cardiovascular reserve, and altered perception of thirst or discomfort.^20^ These physiological changes are compounded by structural and social vulnerabilities, including limited mobility, polypharmacy, dependence on caregivers, and barriers to accessing air conditioning or cooling centers.^1,19,21^ Together, these factors create a profile of heightened susceptibility to heat-related morbidity and, as suggested by our findings, potentially to ADRD.

We did not find differences in the association between heat and ADRD/MCI by sex. Nonetheless, older women have been shown in other studies to experience higher rates both of heat-related mortality and dementia prevalence.^22–24^ Differences in cardiovascular physiology, hormonal factors, and social circumstances, such as living alone or having caregiving responsibilities, may partly explain these trends.^23^ Future research should investigate these dimensions more explicitly, ideally through longitudinal and intersectional analyses that capture both biological sex and gendered social roles.

Our exploratory spline models revealed a non-linear relationship between cumulative heat exposure and dementia risk. While risk increased gradually at first, it rose more sharply beyond approximately 100 days of extreme heat, suggesting a potential threshold beyond which vulnerability accelerates. Although confidence intervals widened at higher exposure levels, indicating reduced precision, the observed pattern suggests that exceeding this threshold may be particularly harmful. Identifying such inflection points could help guide climate-sensitive risk assessments and public health interventions. For example, similar to existing air pollution or UV index warning systems, advisories could be issued for older adults and other at-risk populations once cumulative heat exposure surpasses a defined level linked to elevated risk.

Community-based social determinants of health may influence the relationship between heat and ADRD/MCI. We found that the ADI, a composite measure of neighborhood-level socioeconomic disadvantage, accounted for approximately 9% of the observed association between high heat exposure and ADRD/MCI. In contrast, neither individual educational attainment nor social isolation significantly mediated the relationship. These findings align with a growing literature on the role of the built environment and structural inequities in shaping exposure and resilience to climate-related stressors.^25,26^ Individuals in deprived neighborhoods may have limited access to cooling infrastructure, live in poorly insulated housing, and experience greater exposure to urban heat islands. As such, dementia prevention strategies must integrate climate adaptation measures that prioritize infrastructure improvements in historically marginalized communities.

The findings of this study carry particular relevance for countries and regions facing simultaneous demographic and climate transitions. The confluence of climate change and population ageing is a global phenomenon. Some of the highest projected increases in dementia prevalence are concentrated in regions already experiencing some of the world’s most extreme rises in heat, including the Arabian Gulf and the Indus River Valley, where wet-bulb temperatures near or exceeding 35°C, a theoretical human survivability threshold, have already been recorded.^27^ In this context, dementia risk reduction must be approached not only through clinical or behavioral interventions but also through climate-sensitive public health and urban planning policies.

Several limitations must be considered. First, the cross-sectional design precludes causal inference, and we cannot determine whether heat exposure precedes dementia onset or reflects vulnerability following cognitive decline. However, our use of a large, clinically diagnosed national dataset with detailed covariate adjustments strengthens confidence in the observed association between cumulative heat exposure and cognitive impairment. Future longitudinal studies with incident ADRD/MCI outcomes are needed. Second, while three-digit ZIP code assignment permits national-scale analysis, it may not accurately capture individual heat exposure, which is influenced by occupation, housing, access to cooling, transportation, and daily behaviors. Third, although we adjusted for multiple confounders and conducted mediation analyses, unmeasured variables such as hydration, air conditioning use, or indoor temperatures may introduce residual confounding. Finally, outcome misclassification may be present due to reliance on clinical diagnosis codes, although the use of EHR data suggests strong diagnostic accuracy for ADRD and MCI phenotypes compared to self-report surveys.

Despite these limitations, this study provides new evidence that cumulative exposure to extreme heat is associated with increased odds of ADRD/MCI. As climate change accelerates the frequency and severity of heatwaves, the cognitive health of ageing populations must be prioritized. Our findings suggest that dementia risk frameworks should incorporate environmental heat exposure, alongside air pollution, as part of a comprehensive set of modifiable risk factors. Public health preparedness must include surveillance systems, climate-adapted infrastructure, and early warning mechanisms that specifically account for the needs of older adults and disadvantaged communities. Addressing the dual challenge of dementia and climate change will require integrated, multisectoral approaches that bring together urban planning, environmental science, public health, and gerontology.

## Conclusion

This study provides new evidence that cumulative exposure to extreme heat is associated with increased odds of ADRD and MCI. The strength of the association is greater when exposure is measured using heat index, highlighting the importance of considering humidity in public health risk assessments, and is most pronounced among older adults. Partial mediation by neighborhood-level deprivation underscores the intersection of environmental exposures and social determinants of health, reinforcing the need for targeted interventions. Integrating cognitive health considerations into climate adaptation and public health strategies is essential to protect vulnerable populations and reduce the global burden of dementia. Future dementia prevention frameworks should explicitly include ambient heat exposure alongside established modifiable risk factors.

## Data Availability

All data produced in the present study are available online through the All of Us Researcher Workbench

